# Comparing men who have sex with men and inject drugs and people who inject drugs who are men and have sex with men in San Francisco: Implications for HIV and hepatitis C virus prevention

**DOI:** 10.1101/2021.12.09.21267273

**Authors:** Adelina Artenie, Shelley N. Facente, Sheena Patel, Jack Stone, Jennifer Hecht, Perry Rhodes, Willi McFarland, Erin Wilson, Peter Vickerman, Meghan D. Morris

## Abstract

**Background:** Men who have sex with men (MSM) and people who inject drugs (PWID) carry a disproportionate burden of HIV and hepatitis C virus (HCV) infections. We compared the demographic and risk profiles of MSM who inject drugs (MSM-IDU, i.e., men reached through affiliation with MSM) and PWID who are men and have sex with men (PWID-MSM, i.e., men reached through affiliation with PWID).

**Methods:** We used data from the most recent waves of the National HIV Behavioural Surveillance among MSM (2017) and PWID (2018) in San Francisco. Participants were recruited through venue-based (MSM) and peer-referral (PWID) sampling and completed standardised questionnaires. We compared the characteristics of MSM-IDU and PWID-MSM using bivariate tests.

**Results:** Of 504 participants completing the MSM survey, 6.2% reported past-year injection drug use (MSM-IDU). Among 311 male participants completing the PWID survey, 19.0% reported past-year sex with a male (PWID-MSM). Relative to MSM-IDU, more PWID-MSM were older, identified as bisexual, had lower income, a history of incarceration and were homeless. MSM-IDU had more male sexual partners (median: 10 vs 3) and fewer injected daily (29.0% vs 64.4%) than PWID-MSM. While more PWID-MSM sought sterile equipment from a syringe program (86.4% vs 35.5%), fewer reported using PrEP (15.0% vs 42.9%).

**Conclusion:** The sociodemographic, risk behaviour, and prevention access profiles of MSM-IDU and PWID-MSM in San Francisco suggest that they represent distinct populations who may require tailored HIV and HCV prevention strategies. MSM- and PWID-focused prevention programs should provide combined sexual health and harm reduction messages and services.

## INTRODUCTION

Men who have sex with men (MSM) and people who inject drugs (PWID) are two of the populations at highest risk of HIV and hepatitis C virus (HCV) infection^[1, 2]^. These populations have traditionally been viewed distinctly, with research and interventions generally focusing on either the sexual behaviours of MSM or the injecting behaviours of PWID, despite evidence that that some MSM inject drugs (MSM-IDU) and some male PWID engage in sex with other men (PWID-MSM)^[3-7]^. Studies among MSM have indicated that MSM-IDU have a greater prevalence of HIV and HCV infection, experience more socioeconomic disadvantage and have higher levels of sexual risk behaviours, in addition to also having injection-specific risks compared to MSM who do not inject drugs (MSM non-IDU)^[3-5]^. Conversely, studies among PWID who are men found that, relative to those who do not report sex with other men (PWID non-MSM), PWID-MSM have lower socio-economic stability, are more likely to engage in high-risk injection practices and to be living with HIV^[6, 7]^.

While previous studies have highlighted the distinct characteristics of people with dual risk behaviours relative to other MSM or PWID populations, it remains unclear how MSM-IDU and PWID-MSM compare to each other in their demographic groupings, their drug use and sexual patterns, their primary mode of HIV and HCV infection and how they may be reached. These similarities and differences are important in informing the planning and delivery of HIV and HCV prevention services and the ongoing US national and global campaigns to eliminate these viral infections as public health problems by 2030^[1, 2, 8, 9]^. In the US, the third largest HIV transmission group among men is a joint risk category referred to as “male-to-male sexual contact and injection drug use”^[10]^. Yet, if MSM-IDU and PWID-MSM present distinct risk behaviours, different strategies may be needed to reach each group.

Our primary aim was to characterise similarities and differences between MSM-IDU and PWID-MSM by comparing socio-demographic, injection drug use and sexual patterns, and access to services. As a secondary aim, we also compared the characteristics of MSM-IDU to MSM non-IDU and PWID-MSM to male PWID non-MSM, respectively, to gain a broader understanding of the characteristics of these groups. Our case example is San Francisco, where the burden of HIV and HCV infections is high for MSM and PWID^[11, 12]^, campaigns to eliminate these infections are ongoing^[13, 14]^, and surveys with comparable measures for these populations are available.

## METHODS

We used data collected through US Centers for Disease Control and Prevention (CDC) National HIV Behavioural Surveillance (NHBS) surveys^[15]^, which conducts behavioural surveillance among key populations at risk of HIV infection. We used the most recent data from San Francisco’s MSM (2017) and PWID (2018) surveys. MSM were recruited using time-location sampling and participants were eligible for enrolment if they were ≥18 years, resided in the San Francisco Metropolitan Statistical Area (MSA), and either identified as MSM or had any sex with another man in the previous year^[15]^. The time-location sampling method entailed intercepting MSM at venues where gay and other MSM congregate. PWID were recruited though peer referral and were eligible if ≥18 years, resided in the San Francisco MSA and reported past-year injection drug use^[15]^. The peer-referral method entailed participants referring persons from their social networks who they knew to be PWID.

In both surveys, participants completed an interviewer-administered electronic questionnaire and were tested for HIV. Testing for HCV antibody status was only conducted among PWID. HIV and HCV infection were assessed using standardized testing methods^[15]^. The same core questionnaire was used across both surveys, which collected information about socio-demographic factors, injection drug use, sexual risk behaviours and access to HIV and HCV services. Participants were reimbursed $50 for their time. Both studies were approved by the institutional review boards of the University of California, San Francisco and the CDC.

For this study, we restricted the PWID study sample to participants with self-reported gender as men. Those who reported sex with a man in the past year were categorised as PWID-MSM. Among the MSM study sample, we categorized those who reported injecting drugs in the past year as MSM-IDU. We categorised the remaining groups as PWID non-MSM and MSM non-IDU, respectively.

We used medians and interquartile ranges, and frequency distributions to summarise continuous and categorical variables, respectively. We used Pearson’s chi-square or, alternatively, Fisher’s Exact tests when expected cell counts were ≤5 for categorical variables, and Mann-Whitney U tests for continuous variables to explore differences between groups. All analyses were performed using SAS version 9.4.

## RESULTS

Of the 504 participants completing the MSM survey, 31 (6.2%) were classified as MSM-IDU. Conversely, of 464 participants completing the PWID survey, 311 (67.0%) identified as men and of these, 59 PWID (19.0%) were classified as PWID-MSM. Their characteristics and those of MSM non-IDU and PWID non-MSM are presented in Table 1.

**Table 1:** Socio-demographic, injection drug use and sexual behaviours, and access to services among men who have sex with men and inject drugs (MSM-IDU) or who do not inject drugs (MSM non-IDU) and men who inject drugs and have sex with other men (PWID-MSM) or who do not have sex with other men (PWID non-MSM)

| Characteristic | MSM-IDU<br>(n= 31) | PWID-MSM<br>(N= 59) | p-value† | MSM non-IDU<br>(n= 473) | p-value†† | PWID non-MSM<br>(n= 252) | p-value††† |
| --- | --- | --- | --- | --- | --- | --- | --- |
| **Socio-demographic factors** |  |  |  |  |  |  |  |
| Age |  |  | 0.05 |  | 0.17 |  | 0.14 |
| 18-39 | 20 (64.5%) | 25 (42.4%) |  | 245 (51.8%) |  | 81 (32.1%) |  |
| 40+ | 11 (35.5%) | 34 (57.6%) |  | 228 (48.2%) |  | 171 (67.9%) |  |
| Race/ethnicity |  |  | <0.01 |  | 0.58 |  | 0.34 |
| White | 20 (64.5%) | 23 (39.0%) |  | 238 (50.3%) |  | 122 (48.4%) |  |
| Black/African American | 1 (3.2%) | 12 (20.3%) |  | 26 (5.5%) |  | 52 (20.6%) |  |
| Hispanic or Latino/a/x | 6 (19.4%) | 5 (8.5%) |  | 98 (20.7%) |  | 26 (10.3%) |  |
| Multiple | 3 (9.7%) | 17 (28.8%) |  | 56 (11.8%) |  | 43 (17.1%) |  |
| Other | 1 (3.3%) | 2 (3.4%) |  | 55 (11.6%) |  | 9 (3.6%) |  |
| Sexual identity |  |  | <0.01 |  | <0.01 |  | <0.01 |
| Heterosexual | 4 (12.9%) | 9 (15.3%) |  | 5 (1.1%) |  | 236 (93.7%) |  |
| Gay | 22 (71.0%) | 23 (39.0%) |  | 429 (90.7%) |  | 2 (0.8%) |  |
| Bisexual | 5 (16.1%) | 27 (45.8%) |  | 39 (8.3%) |  | 14 (5.6%) |  |
| Highest level of education completed |  |  | 0.09 |  | <0.01 |  | 0.21 |
| High school or less | 11 (35.5%) | 32 (54.2%) |  | 53 (11.2%) |  | 159 (63.1%) |  |
| Some college, Bachelor's degree and above | 20 (64.5%) | 27 (45.8%) |  | 420 (88.8%) |  | 93 (36.9%) |  |
| Current employment status |  |  | <0.01 |  | 0.06 |  | 0.97 |
| Employed | 17 (54.8%) | 5 (8.5%) |  | 341 (72.1%) |  | 19 (7.5%) |  |
| Unable to work for health reasons | 3 (9.7%) | 21 (35.6%) |  | 16 (3.4%) |  | 89 (35.3%) |  |
| Not employed | 11 (35.5%) | 33 (55.9%) |  | 116 (24.5%) |  | 144 (57.1%) |  |
| Household income |  |  | <0.01 |  | <0.01 |  | 0.83 |
| US\$ 0 - 24,999 | 12 (38.7%) | 49 (83.1%) | | 78 (16.5%) | | 217 (86.1%) | |
| US\$ 25,000 - 49,999 | 5 (16.1%) | 7 (11.9%) | | 92 (19.5%) | | 24 (9.5%) | |
| ≥US\$ 50,000 | 14 (45.2%) | 3 (5.1%) | | 303 (64.1%) | | 11 (4.4%) | |
| Currently homeless | 8 (25.8%) | 39 (66.1%) | <0.01 | 11 (2.3%) | <0.01 | 206 (81.7%) | <0.01 |
| Ever held in detention, jail or prison >24h, past 12 months | 14 (45.2%) | 53 (89.8%) | <0.01 | 73 (15.4%) | <0.01 | 244 (96.8%) | 0.03 |
| **Injection drug use behaviours** |  |  |  |  |  |  |  |
| Age at first injection (Median, IQR) | 30 (23-39) | 22 (16-30) | <0.01 | n/a | n/a | 20 (16-26) | 0.30 |
| Drug most often injected, past 12 months |  |  | 0.53 |  |  |  | <0.01 |
| Meth/amphetamine | 20 (64.5%) | 32 (54.2%) |  | n/a | n/a | 46 (18.3%) |  |
| Heroin | 4 (12.9%) | 13 (22.0%) |  | n/a | n/a | 148 (58.7%) |  |
| Other | 7 (22.6%) | 14 (23.7%) |  | n/a | n/a | 58 (23.0%) |  |
| Injected ≥2 different drug types, past 12 months | 8 (25.8%) | 35 (59.3%) | <0.01 | n/a | n/a | 203 (80.6%) | <0.01 |
| Daily injection, past 12 months | 9 (29.0%) | 38 (64.4%) | <0.01 | n/a | n/a | 204 (81.0%) | <0.01 |
| Used injection equipment previously used by someone else, past 12 months | 8 (25.8%) | 24 (40.7%) | 0.16 | n/a | n/a | 102 (40.5%) | 0.98 |
| Overdose, past 12 months * | 1 (9.1%) | 11 (30.6%) | 0.15 | 0 | 0.30 | 64 (28.1%) | 0.76 |
| **Sexual behaviours** |  |  |  |  |  |  |  |
| Number of male sexual partners, past 12 months (Median, IQR) | 10 (6-20) | 3 (1-10) | <0.01 | 7 (2-20) | 0.38 | n/a | n/a |
| Condomless anal intercourse, past 12 months | 29 (93.6%) | 37 (62.7%) | <0.01 | 381 (80.6%) | 0.07 | n/a | n/a |
| Received money or drugs from a man to have sex, past 12 | 11 (35.5%) | 30 (50.9%) | 0.16 | 26 (5.5%) | <0.01 | n/a | n/a |
| months |  |  |  |  |  |  |  |
| Had a female sex partner, past 12 months | 7 (22.6%) | 30 (50.9%) | <0.01 | 31 (6.6%) | <0.01 | 183 (72.6%) | <0.01 |
| **Testing and use of services** |  |  |  |  |  |  |  |
| Drug treatment, past 12 months | 6 (19.4%) | 18 (30.5%) | 0.26 | 22 (4.7%) | <0.01 | 70 (27.8%) | 0.68 |
| OAT, past 12 months* | 3 (27.3%) | 16 (44.4%) | 0.31 | 0 | 0.02 | 142 (62.3%) | 0.04 |
| Obtained sterile needles from a SSP, past 12 months | 11 (35.5%) | 51 (86.4%) | <0.01 | n/a | n/a | 239 (94.8%) | 0.04 |
| Obtained sterile syringes from a pharmacy, past 12 months | 17 (54.8%) | 29 (49.2%) | 0.61 | n/a | n/a | 92 (36.5%) | 0.07 |
| Tested for HIV, past 12 months** | 19/21 (90.5%) | 30/40 (75.0%) | 0.19 | 304/385 (79.0%) | 0.21 | 159/238 (66.8%) | 0.30 |
| Tested for HCV, past 12 months | 28 (90.3%) | 36 (61.0%) | <0.01 | 258 (54.6%) | <0.01 | 187 (74.2%) | 0.04 |
| PreP awareness** | 18/21 (85.7%) | 29/40 (72.5%) | 0.24 | 373/385 (96.9%) | 0.04 | 115/238 (48.3%) | <0.01 |
| PreP use, past 12 months** | 9/21 (42.9%) | 6/40 (15.0%) | 0.02 | 171/385 (44.4%) | 0.89 | 1/238 (0.4%) | <0.01 |
| HIV positive | 10 (32.3%) | 23 (39.0%) | 0.53 | 87 (18.4%) | 0.06 | 15 (5.9%) | <0.01 |
| HCV antibody positive | n/a | 42 (71.2%) |  | n/a |  | 200 (79.4%) | 0.17 |

Abbreviations: HCV=hepatitis C virus; IQR=interquartile range; SSP= syringe service program; OAT=opioid agonist treatment;
†p-value comparing MSM-IDU to PWID-MSM
†† p-value comparing MSM-IDU to MSM non-IDU
††† p-value comparing PWID-MSM to PWID non-MSM
\*Data available among participants who used opioids in the previous year only
\*\*Data presented among participants who report being HIV-negative
Note: p-values were derived based on Pearson's chi-square test or, alternatively, Fisher's Exact test when expected cell counts were ≤5 for categorical variables, and Mann-Whitney U test for continuous variables

### Comparing MSM-IDU and PWID-MSM

We noted differences between MSM-IDU and PWID-MSM across most socio-demographic measures. PWID-MSM were older than MSM-IDU (57.6% vs 35.5% were ≥40 years), more racially/ethnically diverse (61.0% vs 35.5% identified as non-white) and more identified as bisexual (45.8% vs 16.1%). PWID-MSM were also less educated and fewer were currently employed relative to MSM-IDU, with a majority reporting a household annual income of less than $25,000, experiencing homelessness and having a history of detention.

Injection drug use and sexual behaviours also differed between the two groups. Relative to MSM-IDU, PWID-MSM began to inject drugs earlier (median age: 22 vs 30 years), more injected ≥2 different drugs (59.3% vs 25.8%) and injected daily (64.4% vs 29.0%). Conversely, PWID-MSM reported fewer male sexual partners compared to MSM-IDU (median: 3 vs 10), fewer reported condomless anal sex (62.7% vs 93.6%) and more indicated having a female sex partner (50.9% vs 22.6%). One exception to these differences was the type of drug most often injected, with 64.5% and 54.2% of MSM-IDU and PWID-MSM indicating methamphetamine, respectively.

Some markers of service use differed between the two groups. A greater proportion of PWID-MSM sought sterile syringes from a syringe service program relative to MSM-IDU (86.4% vs 35.5%). Conversely, more MSM-IDU reported using PrEP (42.9% vs 15.0%) and having been tested for HCV (90.3% vs 61.0%) compared to PWID-MSM. The two groups were comparable with respect to HIV testing and opioid agonist treatment. HIV prevalence did not differ between the groups and was high for both MSM-IDU (32.3%) and PWID-MSM (39.0%).

### Comparing MSM-IDU and MSM non-IDU

While most MSM-IDU and MSM non-IDU identified as gay (71% and 91%), a larger proportion of MSM-IDU identified as heterosexual (13% vs 1%) or bisexual (16% vs 8%) compared to the MSM non-IDU group. Key socio-economic characteristics differed between the two groups, with a larger proportion of MSM-IDU reporting having completed only high school education or less (35.5% vs 11.2%), a household annual income of less than $25,000 (38.7% vs 16.5%), current homelessness (25.8% vs 2.3%) and a history of incarceration (45.2% vs 15.4%) compared to MSM non-IDU. We also noted differences for some sexual behaviours. More MSM-IDU received money or drugs in exchange for sex with a man (35.5% vs 5.5%) and having a female sex partner (22.6% vs 6.6%). A larger proportion of MSM-IDU reported prior HCV testing compared to MSM non-IDU (90.3% vs 54.6%). HIV prevalence was also higher among MSM-IDU compared to MSM non-IDU (32.3% vs 18.4%).

### Comparing PWID-MSM and PWID non-MSM

PWID-MSM and PWID non-MSM were comparable on several socio-demographic measures, including age, education, employment and income. A large majority of PWID non-MSM identified as being heterosexual (93.7%), while only 15.3% of PWID-MSM did so. A smaller proportion of PWID-MSM reported current homelessness (66.1% vs 81.7%) compared to PWID non-MSM. Several injection drug use behaviours differed between the two groups. A larger proportion of PWID-MSM reported methamphetamine as their primary drug injected (54.2% vs 18.3%) and a smaller proportion reported injecting ≥2 different drugs (59.3% vs. 80.6%) and injecting daily (64.4% vs. 81.0%) compared to PWID non-MSM. With respect to testing and use of services, we found a larger proportion of PWID-MSM were aware (72.5% vs 48.3%) and had used PrEP (15.0% vs 0.4%) than PWID non-MSM. HIV prevalence was higher among PWID-MSM compared to PWID non-MSM (39.0% vs. 5.9%); no difference was found for HCV prevalence (71.2% vs 79.4%).

## DISCUSSION

Overall, compared to MSM-IDU, PWID-MSM presented greater socio-economic disadvantage on nearly all demographic measures considered, were more likely to be racially/ethnically diverse, and to indicate being bisexual rather than gay. Relative to MSM-IDU, PWID-MSM appeared to have a heavier injection drug use profile but lower sexual risk practices. While MSM-IDU appeared to be more engaged in MSM-oriented prevention programs like PrEP, PWID-MSM were more engaged with syringe service programs, which have been primarily targeted towards PWID. Together, the differences observed in the socio-demographic, risk behaviour and healthcare access profiles of MSM-IDU and PWID-MSM suggest that they represent distinct populations that are present in different social spaces and may require targeted HIV/HCV prevention strategies. More broadly, these findings suggest that harm reduction and healthcare settings catering to MSM and PWID need to reflect the complexity of risk that these groups face, providing a wider range of HIV/HCV prevention messages and services than is suggested by their primary risk behaviour or the label that is ascribed to them.

The extent to which people who engage in both injecting- and sexual-risk behaviours were included in the PWID- or MSM-focused studies could reflect the primary behaviour they associated with and which takes precedence in their day-to-day life or the social and sexual networks they interact with. In a qualitative research study conducted in Denver, some participants reported engaging in male-to-male sex work to sustain their injection drug use patterns, whereas others indicated that injection drug use was only used to enhance male-to-male sex^[16]^. Varying motivations and levels of priority assigned to injecting and sexual practices have also been reported in other studies^[17, 18]^ and explain why some individuals who engage in both behaviours may not identify as MSM or PWID^[16]^. A better understanding of the reasons motivating these practices is needed, as it could increase the extent to which HCV and HIV prevention programs engage with these populations and, ultimately, their impact on minimising risk.

We also noted important differences between MSM-IDU and MSM non-IDU and PWID-MSM and PWID non-MSM, respectively, in line with prior studies conducted among MSM^[3-5]^ and PWID^[6, 7]^. For example, one-third of MSM-IDU indicated receiving money or drugs from a man to have sex, whereas only a minority of MSM non-IDU indicated this practice. In addition, the level of HCV testing was greater in the former group. Conversely, while few PWID-MSM indicated heroin as the most commonly injected drug, more than half of PWID non-MSM did so. Across both MSM and PWID, HIV prevalence was higher among the dual risk groups. Collectively, these findings highlight complex behavioural, service access and HIV infection burden distinctions within both MSM and PWID populations, adding to the importance of providing access to combined sexual health and harm reduction messages rather than targeting specific risk behaviours.

The primary limitation of this study is the small sample size of MSM-IDU and PWID-MSM, which limited the statistical power to detect significant differences between them. It is also possible that we might have incorrectly detected significant differences due to having performed multiple tests. Even so, taken together, the numerous differences observed between MSM-IDU and PWID-MSM across distinct characteristics and measures support the broader finding that these two groups are distinct and should not be conflated with one another. Additionally, aside from HIV and HCV infection status, all other variables were assessed through self-report, which are liable to misclassification errors. However, studies have suggested that self-reported data in these populations are generally valid^[19, 20]^. Finally, the extent to which our findings are generalisable to MSM and PWID populations in other settings is unknown and should be explored in future studies.

In conclusion, our study suggests that MSM-IDU and PWID-MSM represent different populations, with distinct demographic, risk behaviour and healthcare access profiles. In light of ongoing calls to broaden access to HCV and HIV prevention and treatment programs among PWID and MSM to reach 2030 elimination goals, findings indicate a need to provide access to a range of services and prevention messages to both of these populations.

## Data Availability

All data produced in the present study are available upon reasonable request to the authors

